# Exploring Self-Management Barriers and Facilitators Experience by People with Chronic Musculoskeletal Pain - A Qualitative Study

**DOI:** 10.1101/2023.01.18.23284708

**Authors:** Mads Norre Christensen, Simon Kristoffer Johansen, Søren Ravn Sloth, Emil Madsen, Anne Carlsen, Michael Skovdal Rathleff

## Abstract

**Background:** One in three people worldwide will experience chronic musculoskeletal pain (CMP). Due to the long-standing nature of CMP, it is inevitable that people with CMP will have to engage in some form of self-management. As highlighted by multiple clinical practice guidelines, it becomes pivotal that healthcare professionals provide effective self-management support. However, the current effectiveness of self-management interventions seems equivocal. This may be due to self-management interventions not being developed based on an in-depth understanding of the self-management challenges experienced by people with CMP.

**Objectives:** This study explored the everyday barriers and facilitators for self-management experienced by people with CMP currently undergoing rehabilitation in a municipality setting.

**Methods:** We conducted 11 one-time, single-person, qualitative interviews of people living with CMP. The interviews were based on a semi-structured interview guide with open-ended questions and analyzed using an inductive thematic analysis to identify the participants experienced barriers and facilitators for self-management in everyday situations.

**Findings:** Three overarching barriers emerged: 1) biographical disruption, 2) uncertainty and psychological distress, and 3) lack of social support. Further, three overarching facilitators emerged: 1) acceptance and optimism 2) pain-relieving strategies and 3) social support. These barriers and facilitators far exceeded participants capability to control their pain.

**Conclusion:** The majority of the barriers and facilitators concerned the management of cognitive, emotional, social or personal consequences of CMP. Integrating these barriers and facilitators into the development of future self-management interventions may have the potential to improve the effectiveness of these interventions.

**Significance:** Qualitative studies exploring self-management of chronic pain traditionally focus on identification of barriers for pain-management. This study identified several interconnected barriers and facilitators which influenced CMP patients’ everyday self-management. This provided targets for designing self-management interventions to support patients pain-related, psychological, and social self-management practices in everyday contexts.

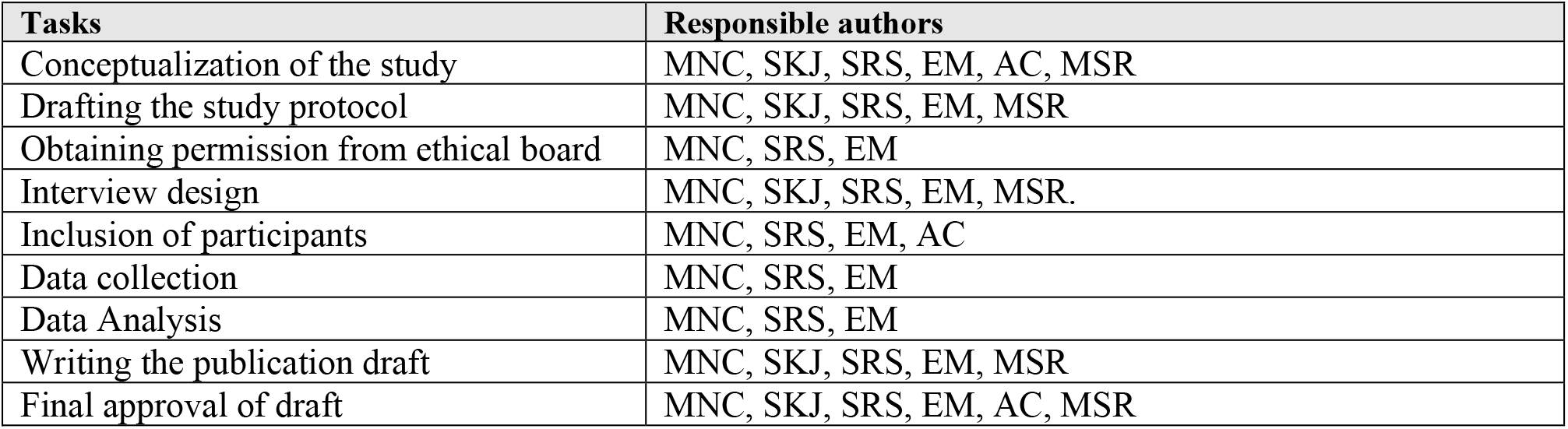

| Tasks | Responsible authors |
| --- | --- |
| Conceptualization of the study | MNC, SKJ, SRS, EM, AC, MSR |
| Drafting the study protocol | MNC, SKJ, SRS, EM, MSR |
| Obtaining permission from ethical board | MNC, SRS, EM |
| Interview design | MNC, SKJ, SRS, EM, MSR. |
| Inclusion of participants | MNC, SRS, EM, AC |
| Data collection | MNC, SRS, EM |
| Data Analysis | MNC, SRS, EM |
| Writing the publication draft | MNC, SKJ, SRS, EM, MSR |
| Final approval of draft | MNC, SKJ, SRS, EM, AC, MSR |

## 1. Introduction

Experiencing pain is a normal part of life and an essential function for learning to react on or avoid potential threats to the body. For the majority of people, a painful experience will resolve as the underlying cause or potential threat ceases. However, in the 1.71 billion people worldwide living with a painful musculoskeletal condition, up to one in every three will end up developing chronic musculoskeletal pain (CMP) (Wu et al., 2020,Stevans et al., 2021,Itz et al., 2013,da C Menezes Costa et al., 2012). To these people, CMP can become a major barrier for living meaningful lives in accordance with their personal values, as CMP is associated with significantly reduced physical functioning, social participation, quality of life and psychological wellbeing (Mills et al., 2019,Dueñas et al., 2016,Cáceres-Matos et al., 2020) Further, people with CMP have higher healthcare use, sickness absenteeism and loss of productivity (Mills et al., 2019,Dueñas et al., 2016,Cáceres-Matos et al., 2020,McDonald et al., 2011). Identifying effective treatment options are therefore important from both a societal and individual perspective. International clinical guidelines and systematic reviews of randomized controlled trials recommend the use of non-pharmacological approaches, such as patient education, exercise therapy and psychologically-informed therapies to provide self-management support for a variety of musculoskeletal conditions (Lin et al., 2020,Babatunde et al., 2017,National Institute for Health and Care Excellence, (NICE), 2021). Focusing on self-management support is pivotal, as people with CMP inevitably will have to self-manage their chronic condition, as well as the related biopsychosocial consequences, in the timespan when they are not together with a healthcare professional (Van de Velde et al., 2019,Lorig and Holman, 2003). Therefore, healthcare professionals should prioritize providing people with CMP with effective self-management support aimed at improving the management competencies of people with CMP outside the context of healthcare, as this is where the daily self-management process occurs (Lorig and Holman, 2003,Corbin, Juliet and Strauss, 1985,Corbin, Juliet M. and Strauss, 1988). However, the current evidence concerning the effect of self-management interventions for CMP have only shown small and often clinically insignificant effects (Du et al., 2011,Du et al., 2017,Elbers et al., 2018,Oliveira et al., 2012,Geraghty et al., 2021). This could be due to how self-management interventions for people with CMP rarely are developed based on an in-depth understanding of the barriers and facilitators for self-management encountered by people with CMP in their everyday lives. Thus, current self-management interventions may be limited in their effectiveness as they may not address context-specific biopsychosocial domains of importance for people with CMP, when entering into rehabilitative treatment for their CMP (Lorig and Holman, 2003). Currently, there is a dearth of studies exploring the relationship between the individual and contextual barriers and facilitators for self-management experienced by people with CMP in the context of receiving rehabilitative treatment. The aim of this study becomes to explore the everyday barriers and facilitators for self-management experienced by people receiving rehabilitative treatment for CMP, to identify principles for enhancing the quality of treatments and management education for people living with CMP on a broader scale.

## 2. Methods

This qualitative study used one-time, single-person, semi-structured, qualitative interviews informed by the guidelines of Kvale & Brinkman, and based on a phenomenological approach, as described by Zahavi (Zahavi, 2003,Liamputtong Pranee, 2019,Kvale and Brinkmann, 2009) Ethical approval was sought from the Regional ethical committee of Northeren Jutland, which waived requirement of full ethical approval due to the non-interventional nature of the study, in accordance with national guidelines. All participants received both written and oral information about the study details, data management and their participatory rights, and informed consent was obtained from all participants prior to their enrollment as required in the GDPR.

### 2.1 Recruitment and setting

The participants were initially recruited through two rehabilitation centers in the Northern Denmark Region, where they currently engaged in treatment. They were screened according to the criteria in table 1 by one of the authors (AC). If the participants fulfilled the criteria, their contact information were passed on to the main three investigators (MNC, SRS, EM). Through telephone, a secondary screening was conducted. If the participants still fulfilled the eligibility criteria, an appointment for the interview was arranged. To participate in the study, the participants had to fulfill the following eligibility criteria:

**Table 1:** Eligibility criteria

| Inclusion criteria | Exclusion criteria |
| --- | --- |
| • Individuals experiencing chronic musculoskeletal pain in one or more anatomical regions such as muscles, tendons, joints or bones, that: 1) had persisted or recurred for a period of longer than 3 months, and 2) was associated with significant emotional distress (e.g. anxiety, anger, frustration, or depressed mood) and/or significant functional disability (e.g. interference with activities of daily living and participation in social activities)<sup>25,26</sup>.<br>• Individuals between 18-67 years of age.<br>• Individuals able to speak and understand the Danish language. | • Individuals with chronic pain explained by a specific diagnosis or cause such as an infectious, immunological, neurological or rheumatological disorder<sup>25,26</sup>.<br>• Individuals with cognitive or psychological problems compromising their memory or ability to independently explain their own medical history and experience. |

### 2.2 Developing the interview guide

The interviews were based on an inductive and semi-structured interview guide containing open-ended questions regarding the participants experienced barriers and facilitators for self-managing their CMP in everyday contexts (Kvale and Brinkmann, 2009) *(Appendix 1)* This approach was chosen to avoid any predetermined foci of interest, as well as ensuring that the participants had the possibility to articulate any topic of importance to them. The interview guide was designed by the main investigators (MNC, SRS, EM) with inputs from (MSR & SKJ). The final interview guide contained a total of four overarching interview questions concerning the participants perceived barriers and facilitators, and several open-ended probing question, which the interviewer could use to facilitate elaboration of the participants experiences. Before the initial interviews, three pilot-interviews were conducted to ensure that the participants understood the question formulation, and the phenomena was examined in an appropriate and comprehensive manner. Based on feedback from these pilot-interviews the final adjustments of the interview guide were made.

### 2.3 Qualitative interviews

The qualitative interviews were conducted by the 3 main investigators (MNC, SRS, EM) at one of the rehabilitation centers that the participants were already accustomed to and visiting on a regular basis. The interviews lasted approximately 60-90 minutes each. Throughout the interviews, two of the main investigators were present (1 main interviewer, 1 observer) to ensure that the main interviewer addressed all topics of relevance. Furthermore, the main investigators were aware of both their verbal and non-verbal communication throughout the interviews, while seeking to create a trustful and empathic relationship with the participants and cultivate a safe environment for the participants to articulate their experiences within (O’Keeffe et al., 2016). The interviews were recorded using an Olympus WS-553 dictaphone and transcribed in Microsoft Word version 14.46.

### 2.4 Data analysis

The qualitative analysis was conducted using the NVivo software. We used thematic analysis as described by Braun & Clark (Braun and Clarke, 2006) The thematic analysis approach was used to inductively identify themes related to participants experienced barriers and facilitators for managing their CMP. Each of the six phases in the thematic analysis was conducted through collaboration between the three main investigators (MNC, SRS, EM), and agreement was sought during each phase to ensure credibility of the findings. In the first phase, all investigators read through the transcripts to familiarize themselves with the totality of the data set. In the second phase, transcripts were preliminarily coded to identify statements, which potentially could constitute a barrier or facilitator for self-management, and statements expanding the meaning of potential barriers and facilitators. In the third phase, an in-depth analysis of the preliminary codes was conducted to organize the codes into potential themes and sub-themes. In the fourth phase it was checked if a coherent thematic pattern could be identified across the dataset. Secondly, the themes were reviewed and refined externally in relation to the totality of the dataset to ensure the themes reflected the meanings of the participants in an accurate and valid manner. At the end of the fourth phase, a thematic map was developed to establish an overview of the thematic analysis and ensure full transparency (*Appendix 2)*. In the fifth phase, the themes were clearly defined, and the essence of each theme were described by extracting and analyzing the meaning of each code within the themes. The sixth phase consisted of the final reporting of the themes.

## 3. Results

### 3.1 Sample description

We conducted a total of 11 semi-structured interviews with no new significant themes emerging after the 9^th^ interview. Participants were Danish speaking adults ranged in age from 19-67 (median = 56 years), and the majority of the participants were women (73%). The participants had experienced CMP in a period ranging from 10 months to 39 years (median = 48 months), and had consulted several different healthcare professionals, as well as attempted several different treatment options throughout their course of treatment *(Table 2)*. All of the participants were currently engaged in a course of treatment at one of two rehabilitation centers in the Northern Jutland area.

**Table 2:**
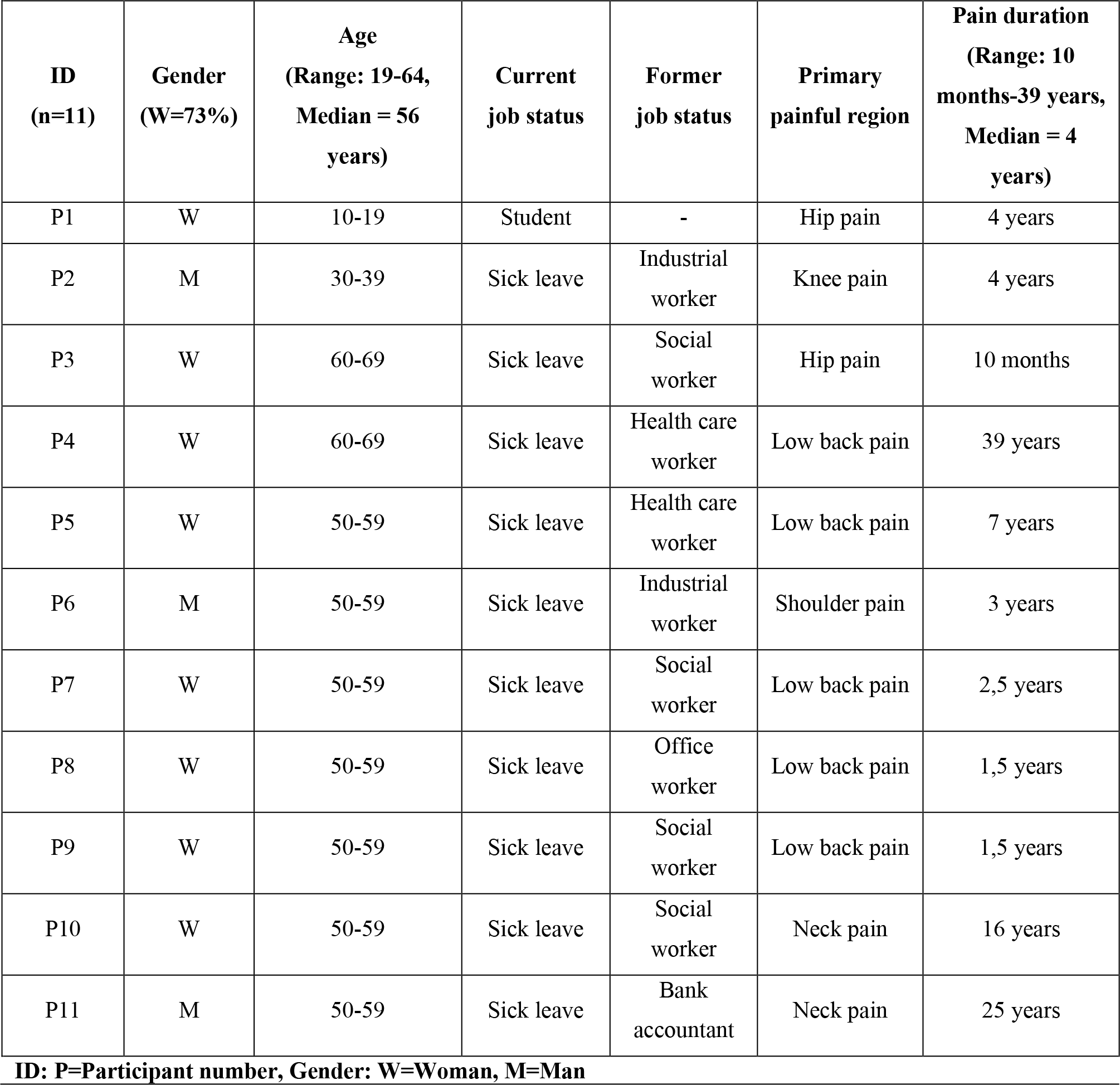
An overview of the eligibility criteria applied during the inclusion of participants. Clinical characteristics of participants. Characteristics of the participants (n=11) included in this study.

### 3.2 Interpretation of themes

As a result of the thematic analysis, we identified a total six storybook themes. Of these, three themes were classified as barriers for self-management, and three were classified as facilitators (See figure 1). The six main themes are presented as parts of a shared narrative describing different aspects of the participants shared experience with managing their CMP.

**Figure 1:**
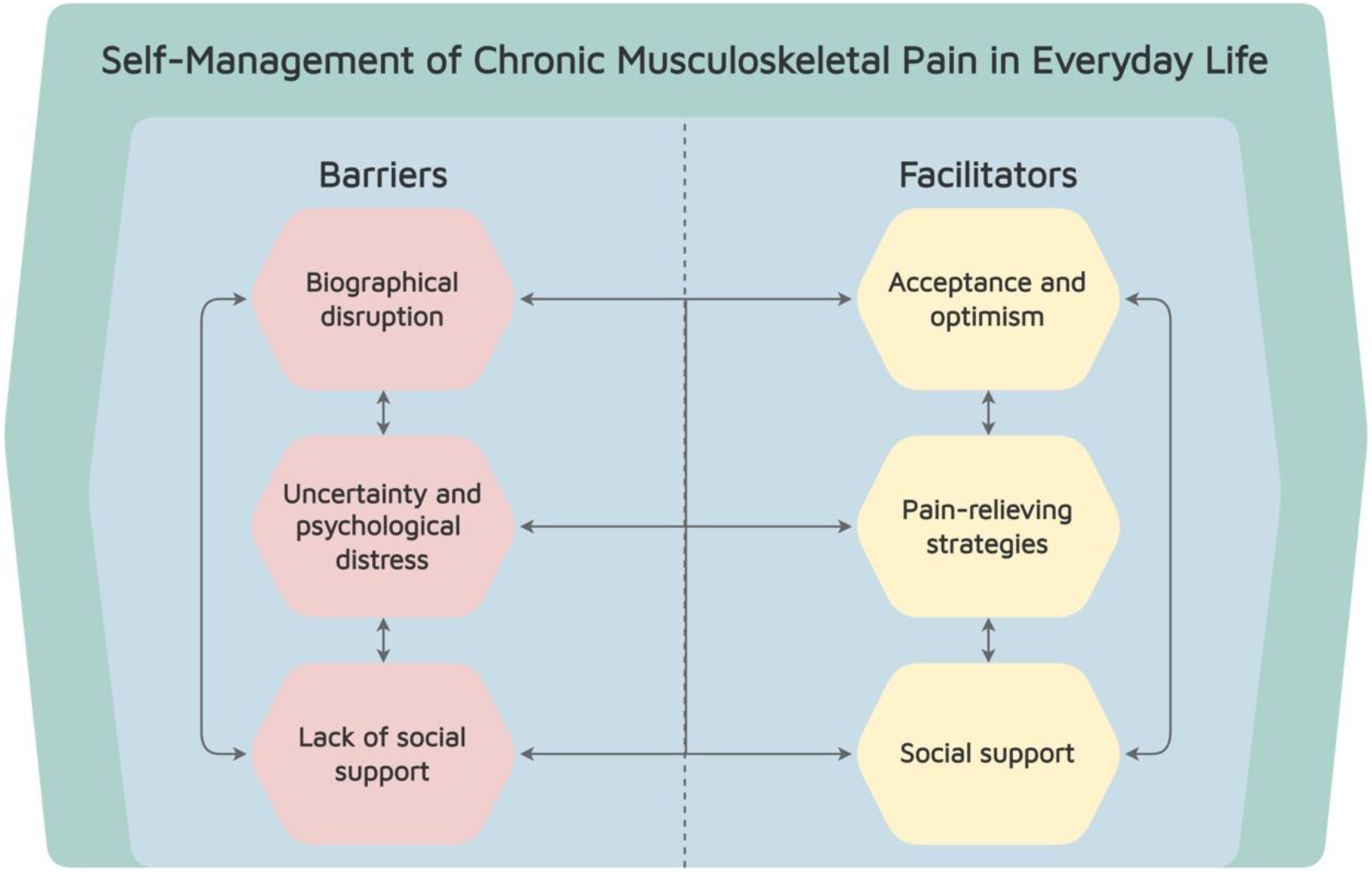
A conceptual model illustrating the relationships between Barriers and Facilitators of self-management of CMP identified during the analysis of CMP patients’ descriptions.

### 3.3 Barriers for self-management

The participants reported a range of interrelated barriers, which made it difficult to self-manage their CMP on a day-to-day basis. Based on the analysis, three overarching themes emerged: 1) biographical disruption, 2) uncertainty and psychological distress, and 3) lack of social support.

### 3.4 Theme 1 Biographical disruption

The majority of the participants described that it was difficult to self-manage their CMP, because they, due to the risk of painful flare-ups and fatigue, felt a need to choose between equally meaningful activities at home. As such, they often felt that their identity, and personal values were threatened, and felt confronted with a need change themselves and their everyday lives. Many participants described having experienced difficulties finding acceptance of their new life situation and had sought out different healthcare professionals hoping they were able to find a treatment approach that could remove their pain, so they could become themselves again.

> *P8: “The hardest part for me is definitely the mental part. Learning to accept that everything is, as it is, and that you’re no longer able to do the things, that you once were able to do, is so tough. I still haven’t found the acceptance of not being able to do the things that I could two years ago. I’ve never been that type of person, who has asked others for help, irrespective if it was to set up a lamp or change a power socket. But now there are just so many things, that I’m no longer able to do on my own*.*”*

Furthermore, many participants explained that they felt a need to give up on social roles (e.g. as a parent, child, friend or colleague), because they were no longer able to participate in social activities, or contribute as much as usual to physical tasks. This daily task of choosing between equally meaningful activities was described as a difficult and iterative learning process, which the participants were necessitated to go through, if they wanted to become able to get through their everyday lives without experiencing flare-ups or becoming mentally and physically exhausted. However, it also made it difficult for the participants to manage their daily resources, as it required them to make significant changes in their priorities, identities, social roles, values and everyday lives.

> *P7: “It is very difficult to manage your daily resources. Because of my pain, I have less energy than a normal person, but the same amount of tasks to do. I usually prioritize basic household activities such as cleaning and my personal hygiene, and some of the things I need to do every day to feel good are to exercise and be physically active. This is actually on top of my priority list for spending the energy, which I have available per day, but this also means, that you have to say goodbye to other things, and often this ends up being social activities. If I’m tired and I’ve spend all my energy, then I’m not able to go see someone or have visits, because the pain is so mentally exhausting. So, I constantly have to plan and manage how I spend my energy*.*”*

Many of the participants described feeling like strangers in their own bodies, and either had lost, or were afraid of losing, their feeling of autonomy. As such, the participants often found it difficult to accept the need to receive help from their friends, families or healthcare professionals in order to self-manage. Further, many of the participants told they experienced a lack of self-worth, because they had been forced to give up on the personal values and social roles, which contributed to their identity and self-confidence.

> *P5: “I feel like a poor person. Someone who is less worthy, because the things that made me very worthwhile, was that I was really good at my job, and that I could take care of everything and everyone, whom I had to take care of. Everything in my life used to be under control. Now my husband is doing our grocery shopping, and I can’t even help him by lifting the grocery bags. That is really frustrating. From being the person who looked after everyone and took care of our home, to who I am now, makes me feel less worthy. All the roles I had have disappeared from my life*.*”*

### 3.5 Theme 2 Uncertainty and psychological distress

The majority of the participants described that a lack of understanding about the cause of their pain, how to manage it, and what to expect of their future, made it difficult for them to self-manage the negative impact that CMP had on their everyday lives. Further, several participants described how it was difficult to figure out which information to believe in, as they often received contradicting information from healthcare professionals. As such, the participants expressed having experienced uncertainty and worries on why they experienced pain, challenges about how to self-manage on a day-to-day basis, and difficulty explaining the severity of their situation to friends, families, colleagues or employers.

> *P1: “I am that type of person, who likes to be informed about my condition, so I can make a plan according to that. I think it would make my whole situation a lot easier to handle, if I understood why I am in pain. So, I actually don’t feel like I am able to move on, before I know what I’m going through. Then I would at least be able to prepare myself for the things to come and make a plan for the future. But this state of uncertainty that I’m currently in, is so unpleasant, and it frightens me when I don’t understand why I’m in pain*.*”*

The majority of the participants explained that their lacking understanding, in addition to their lack of acceptance, often led to negative thoughts and emotions, such as anxiety, worry, catastrophizing, depressed mood, poor expectations and emotional distress. As an example, many participants described that they were afraid they would never get better and felt they would never become themselves again.

> *P8: “It is just as important to understand your pain, as it is to accept it. If just I could get a proper explanation from a healthcare professional about why I am in pain, why it is the way it is, and why nothing can be done to make it better. Then hopefully all my speculations and worries would stop, so I can start concentrating my energy on some of the other challenges I need to tackle. That would make my life a lot easier. My head is completely filled with speculations about my pain and worries about what is going to happen in the future*.*”*

The presence of negative thoughts and emotions were by the participants experienced as being both physically and mentally exhausting, which made it even more difficult to find the energy to self-manage during their everyday lives. Further, the negative thoughts and emotions were often described as interfering with the participants ability to sleep, leading to even less physical and mental energy.

> *P10: “I’m constantly having all these thoughts about how I’m going to manage everything. When I finally go to bed at night, I’m having a really hard time falling asleep. Actually, I can’t even recall the last time that I slept whole a night. Typically, I’m awake every 1-2 hours each night. This is partly because of my nightly aches and pains, but also because I’m filled with speculations and worries. The fact that I don’t get close to six hours of continuous sleep really affects me. And it has a huge impact on my mental and physical energy the following day. So I have to sleep during the day sometimes. It’s like a vicious circle that is hard to break*.*”*

Many of the participants also told that they experienced a constant feeling of guilt and shame, because they saw themselves as a burden to their families, friends and colleges due to their lack of ability to contribute, participate or help as much as they used to. For several of the participants, this feeling resulted in social isolation, as they thought it was better for their friends, if they were not there.

> *P2: “I feel like I would be a burden to them. I am not able to participate in all the things that we could do in the past. Water skiing and stuff like that. We were very active in the group of friends I was a part of. So, I will just become a limitation to them. If I am there, I know they will say no to some of the things they would like to do. Why should it affect them, that I have been injured? I know it’s wrong, because they don’t think that way, but I just feel that it is best for them*.*”*

### 3.6 Theme 3 Lack of social support

The majority of the participants described that it was difficult to self-manage, because their friends, family, colleges and healthcare professionals did not believe in their invisible and painful condition. Lack of trust, social support and responsiveness from their families, friends, colleagues, employers and healthcare professionals made the participants feel helpless and facilitated a feeling of wanting to give up during their course of treatment.

> *P8: “Other people constantly think they know so much about why you’re in pain, and what you should do about it. Like I haven’t tried everything! So, as well as it is hard for yourself to accept your condition, it’s also difficult for others to understand and accept it. I think that is a major challenge, because it’s really unpleasant and affects mood in a negative way, when other people don’t believe in you*.*”*

Furthermore, the lack of trust and responsiveness from healthcare professionals led to a feeling of not being validated and understood, which by the participants were perceived as a major barrier to establish trustful relationships with their healthcare professionals, in which they were able to receive the necessary support enabling them start self-managing their condition.

> *P2: “The lack of help and responsiveness from healthcare professionals has left me with a feeling of helplessness. (*..*) So, I’m quite angry at them, and I’m having a hard time trusting healthcare professionals, because I thought they were there to help me. When you experience that isn’t how it is, then you become distrustful. And you start feeling that you’re not able to get the help that you need. It’s ridiculous they won’t help a young person like me. Maybe things had not come this far, if I had been able to get some help – but I couldn’t*.*”*

### 3.7 Facilitators for self-management

The participants also reported a variety of facilitators, which improved their ability to self-manage their CMP on a day-to-day basis. Based on the analysis, these insights were categorized into three overarching themes: 1) acceptance and optimism 2) pain-relieving strategies and 3) social support.

### 3.8 Theme 4 Acceptance and optimism

The majority of the participants explained that it became a lot easier to self-manage their CMP, when they learned to accept their situation and limitations. Acceptance was described as a prolonged, continuous and difficult process, which was accomplished through an increased understanding of their pain, as well as support from the participants families, friends and healthcare professionals. Further, attaining acceptance was described as an indispensable attribute, which enabled the participants to stop confronting themselves with things they were no longer capable of doing. As such, acceptance improved the participants ability to maintain a positive mindset and focus on possibilities instead of barriers and challenges, and on how they could begin self-managing their situation in the best possible way. Hence, acceptance was described as a crucial starting point for learning to self-manage their CMP, as it was impossible to self-manage a condition, which the participants did not accept the presence of.

> *P7: “Once you learn to accept your limitations, it becomes a lot easier to learn to manage your condition. It becomes much easier to enjoy what is left, and it helps you to stop confronting yourself with all the things, that you can’t do anymore. But acceptance is an unbelievably difficult process to go through, and how long it takes is very individual and varies from person to person. I see it as different phases, that you need to go through. You need to acknowledge your limitations, accept them and then try to move on. Each phase is unbelievably hard to go through, and it takes a lot of time. But I’ve actually gotten into the ‘move on’-phase. I do, however, go back to the ‘acceptance’-phase once in a while. It takes many years to acknowledge and accept your limitations. Because you have to change your entire life. It gets better with time though. You just have to take one day at a time*.*”*

Acceptance were often described synonymously in terms with maintaining positive expectations, optimism and being perseverant, which were also experienced as essential psychological traits, if the participants should succeed in not giving up during the course of their treatment. Thus, finding acceptance, as well as being able to cope with adversity in a positive way, were by the participants perceived as crucial for counteracting the cognitive, emotional and personal challenges of their CMP.

> *P9: “Keeping a positive mindset helps you being optimistic, hope for a better future and to move on. I think that is what’s keeps me going. If I were to sit all day and feel sorry for myself while thinking about my pain all the time, then I might just as well give up. But when you’re having something to look forward to, it makes you want to keep on fighting. So, it’s important to focus on all the positive things in your life”*.

Further, the participants described that the experience of having successes, as well as receiving encouragement and positive feedback from their friends, families, and in particular healthcare professionals, were a huge motivation to keep on fighting and maintain a positive mindset despite being confronted with challenges and adversity during their everyday lives.

> *P3: “Recently, I’ve it experienced that I’ve started to feel slightly better, and it really helps me to think, that of course there is hope. Previously I thought that I was going to feel like this for the rest of my life, but I’ve actually gotten my hope back*.*”*

### 3.9 Theme 5 Pain-relieving strategies

All the participants described that experiencing pain-relief facilitated their ability to self-manage their CMP, as it gave them a feeling of hope for a better future and led to more physical and mental energy. Further, some of the participants told that experiencing pain-relief were able to counteract some of their negative thoughts and emotions. Pain-relief could stem from almost any kind of treatment approach, such as physical activity, exercise, acupuncture, manual therapy, surgery, injections, electrotherapies, mindfulness or analgesics. However, the participants experienced a significant ambivalence towards the use of analgesics, because they were concerned of any harmful side effects.

> *P9: “I really like having some exercises that I can do at home to relieve my pain. It makes me feel like I’m doing something good for myself, and that I’m doing an effort to get better. I don’t want to just sit down waiting for some other person to solve my problem. It means a lot to me, that I’m able to make an effort myself to get better. And I actually think, that it helps me a lot both physically and mentally by maintaining a hope for a better future and increasing my daily energy*.*”*

Of the different pain-relieving strategies, the majority of participants had a particular preference for learning to self-manage their daily resources through the use of pacing strategies, such as planning ahead and incorporating activity breaks. Integrating these strategies enabled the participants to participate in a larger amount of valued or social activities and increased their ability to contribute during activities of daily living without experiencing a painful flare-up and becoming mentally or physically exhausted.

> *P8: “I’m not going to do anything, that I haven’t planned ahead. Because you might end up spending your daily energy on unnecessary stuff. So, for me, personally, planning ahead has been crucial, and I usually plan everything in detail. At first, it was really difficult to do, but with help from my physiotherapist it has become a lot easier. I’ve learned to accept my limitations, and explain myself to other people, so they know how I feel. Before, I used to just vacuum my whole apartment at once, and then feel terrible for the two following days, but now I’ve learned to integrate breaks and split daily tasks into smaller bouts. At least I’m getting something done now*.*”*

### 3.10 Theme 6 Social support

All the participants described that emotional support from their families, friends, colleges, and healthcare professionals were pivotal for their ability to self-manage the negative impact CMP on their everyday lives. Feeling understood, validated and able to share their daily challenges and emotional frustrations with their friends and families, had a significant impact on the participants abilities to accept their situation, and manage the occurrence of negative thoughts and emotions.

> *P1: “I think the biggest help for me, has been the understanding from my family. Having someone close to you every day, who wants to listen to you means a lot. It has made this whole situation a lot easier for me to handle. And it makes you feel like you’re not in this alone, as you’re always within a safe environment, where your family is there to help you with your struggles. They can give you some advice, and when you’re having a flare up, it’s just really nice to have someone to talk to*.*”*

In addition, the majority of the participants explained that physical support with activities of daily living from their friends and families were indispensable, as it helped them physically and mentally to get through challenges with activities of daily living. Spending time with their friends and families were by many of the participants also described as having a positive impact on their mood and pain interference, as it reduced their attentional bias towards their pain.

> *P7: “Being together with your family and friends improves your mood, and an improved mood makes your pain a lot better. If we’re going back to the social part, I’ve actually also experienced something else. Being together with someone actually helps me to shift my attention away from my pain and onto the conversation instead. When I’m going for a walk on my own and my pain starts to flare-up it’s really difficult to shift my attention away, but that’s not a problem, when I’m together with someone else*.*”*

Most of the participants also described that being acknowledged, listened to and having their pain validated by healthcare professionals, were a crucial first step for establishing strong and trustful relationships with their healthcare professionals. Establishing such relationships were experienced as a major facilitator of acceptance, as well as their feeling of having the necessitated support to start learning how they self-manage their condition.

> *P3: “Talking with my physiotherapist has helped me a lot. Before I started at the physio, I used to tell myself I was just being silly. I will never forget this one time, when she took my hand, looked at me, and said: “Stop telling yourself that you’re just being silly all the time. I see that you’re in pain. It’s not just something going on inside your head”. It meant so much to me. To be acknowledged, understood, and to know I’m in a good place has helped me so much. To have a healthcare professional who believes in you, and who is able to explain to you why you’re in pain, has helped me understand that there’s actually a reason why I’m in pain”*.

### 3.11 Summary of findings

The findings from the analysis were summarized in a conceptual model, which outlined the underlying relationships between the six themes. The model was designed to illustrate how participants management of CMP was a complex ongoing activity, and how the barriers and facilitators were organized in two clusters of behavioral outcomes, which acted as each other’s opposites and influenced the participants ability to self-manage in everyday situations.

The themes in each cluster were distributed across the biological, psychological, and social-contextual domains, to signify how people with CMP’s challenges with self-management extended beyond pain management. Read from the right, the model illustrated how *‘biographical disruption’, ‘uncertainty and emotional distress’* and ‘*lack of social support’* formed a cluster of interconnected barriers, which inhibited participants self-management ability. Furthermore, by illustrating the themes as interconnected, the model illustrated how experiences in one domain (i.e. the social domain) could ripple into other domains. Taking point in this mechanism, the model illustrated how *‘acceptance’* (top right) acted as a base or gateway for changing people with CMP perspectives on their management abilities and using their management experiences to develop *‘pain management strategies’* (middle) and utilizing *‘social support’* (bottom right). Taking points in this progression, the model identified *‘acceptance’*, enhancing ‘*pain management strategies’* and garnering *‘social support’* as targets for teaching people with CMP how to explore and progress their self-management in everyday situations. This argues for expanding the scope of treatments, towards adapting a biopsychosocial perspective, with equal focus on barrier identification and supporting people with CMP’s pain-related, psychological and socio-contextual self-management according to people with CMP’s experienced needs for support, to ensure that they gain positive behavioral outcomes which enable them to maintain their efforts towards self-management in everyday situations.

## 4. Discussion

Our findings revealed that the experienced of barriers and facilitators for self-management extended beyond participants capability to control their pain. Only one of the identified barriers and facilitators concerned pain-relieving strategies, while the remaining concerned the cognitive, emotional, social, dimensions of CMP patient’s everyday self-management. This insight has several implications which should be considered when designing future management interventions for people with CMP.

### 4.1 Explanation of findings

Our results align with the ongoing shift in the understanding of self-management, which highlights that self-management encompasses multiple biopsychosocial challenges experienced by people with CMP^13,29^. Our findings expanded upon this by illustrating how self-management should be understood as a daily, complex, and continuous process primarily taking place outside healthcare settings, in the everyday life of people with CMP (Lorig and Holman, 2003,Bodenheimer et al., 2002,Townsend et al., 2006,Van de Velde et al., 2019) A key insight was how identified barriers and facilitators for self-management were interconnected and interacted with each other, as described by Strauss and Corbin in their three lines of self-management (Corbin and Strauss, 1985). This insight challenged the premise of how learning to manage CMP didn’t follow a predictable trajectory, as its often described in self-management literature (Lorig and Holman, 2003). Rather, the relationships indicated that participants’ perspectives on their CMP shifted, thus influencing people with CMP’s ability to purposefully apply strategies like pacing or identifying pain thresholds to avoid flair-ups and fatigue (Paterson, 2001,Khanom et al., 2020). From the identified barriers, participants highlighted how internal phenomena like uncertainty, lack of acceptance, negative thoughts and insomnia were interconnected and gradually impacted participants ability to assert control of their CMP (Bandura, 1977). In contrast, acceptance was described as a major facilitator that enabled participants to stop ruminating, counteract negative emotions, and commence exploring ways of managing their CMP. This aligns with previous research highlighting acceptance as an essential component in participants efforts towards learning to self-manage, as it allowed people with CMP to adjust their pain beliefs, performance, and management practices towards maintaining balance with their CMP (Devan et al., 2018,Toye et al., 2021,Lennox Thompson et al., 2020,Lachapelle et al., 2008,Kostova et al., 2014). Therefore, helping people with CMP to accept their CMP and limitations it imposed on them was important and should be prioritized to empower self-management between consultations.

### 4.2 External support, encouragement, and the locus shift

Another key finding related to how external factors like validation and social support impacted participants ability to make everyday management decisions. Participants described how an absence of validation and responsiveness form social relationships and healthcare providers constituted a management barrier, related to the emergence of negative thoughts, emotions, and decreased willingness to explore new management strategies. In contrast, physical and emotional support from social relations, validation and responsiveness were described as efficacy information, which acted as a facilitator for improving people with CMP’s ability to cope and explore managing their CMP at home (Bandura, 1977). Self-management literature highlights how contextual factors may impact people with CMP’s ability to apply advice or instructions from clinicians to manage actual or potential impacts of disease at home (Kongsted et al., 2021). Nevertheless, our findings expanded upon this by illustrating how external factors like trust and support facilitated exploration, while unsupportive environments fostered perseverance and conformity to maintain existing roles and value systems, as documented by Townsend et al. (Townsend et al., 2006). These findings align with Bodenheimer et al.’s collaborative care principles, where the health professional assumes the role as the expert in disease and management, whereas people with CMP acted as experts in their unique life challenges and integrating self-management into everyday activities (Bodenheimer et al., 2002). Our findings are consistent with several other studies that have investigated barriers and facilitators of people with CMP. However, these studies primarily focus on people with CMP taking part in a specific self-management interventions during research projects (Spink et al., 2021,Devan et al., 2018,Bair et al., 2009,Franklin et al., 2016,Gordon et al., 2017). Thus, previous studies focus on the clinical encounter as the driver for behavioral change, and negate to reflect upon the formative effects of navigating everyday contexts via trail and error (Corbin and Strauss, 1988,Corbin and Strauss, 1985). Previously identified barriers for self-management include external factors, like financial constraints, lack of social support and accessibility related to participation in self-management programs (Spink et al., 2021,Devan et al., 2018,Gordon et al., 2017). Still the studies focus on barrier identification meant that the question of how to support people with CMP and clinicians’ health information management, to ensure that self-management- band social support delivered is applicable and easily integrated into the healthcare specialists’ treatments and personal ecologies of people with CMP alike (Moen and Brennan, 2005). Previous studies highlight support groups as facilitators for integrating self-management between consultations, as people with CMP can provide and gain corrective experiences from peers (Spink et al., 2021,Devan et al., 2018). While support groups were not specifically mentioned in this project, this finding corresponded well with social support being a facilitator of self-management in this current study.

### 4.4 Clinical implications

In the current study, the majority of barriers and facilitators for self-management concerned the personal, cognitive, emotional, social or personal consequences of living with CMP. This might constitute a major challenge, as several studies have shown that many clinicians, while recognizing the need for a biopsychosocial approach in managing CMP, prefer dealing with the more biomedical aspects of CMP, as they feel unprepared, uncomfortable and not adequately trained to address and manage the multidimensional aspects of CMP. (Synnott et al., 2015,Alexanders et al., 2015,Driver et al., 2017,Ng et al., 2021,Holopainen et al., 2020) Further, this might constitute a major challenge for many clinicians, as it indicates that the beliefs and assumptions are still entrenched within a biomedical and expert-centered model, which contradicts the biopsychosocial and person-centered approach deemed necessary for effective self-management support based on this study’s findings. The result is that people with CMP are left to struggle with the complex task of applying and integrating management advice, whilst navigating the psycho-social impacts of CMP between consultations on their own (Moen and Brennan, 2005). Thus, our findings support the need for a paradigm shift in healthcare towards a more biopsychosocially-orientated and person-centered approach. Further, our results highlight that people with CMP perceive a wide range of pain-relieving strategies as facilitators for self-management. However, as self-management takes place outside the healthcare, clinicians might in particular benefit from centering their treatment approaches around active self-management strategies, such as teaching pacing, graded exposure/activity, physical activity or exercise, as these are accessible without the presence of clinicians (Geneen et al., 2017,Hayden et al., 2021,López-de-Uralde-Villanueva et al., 2016,Macedo et al., 2010,Guy et al., 2019)

### 4.5 Limitations

A challenge when interviewing participants with pain, relates to participants’ abilities to recall pain experiences and attribute meaning to them due to recall bias or saliency (Gendreau et al., 2003) Secondly, the stigma surrounding living with chronic pain could increase social-desirability bias, leading participants to over- or underreporting certain aspects of their CMP experience (Bergen and Labonté, 2020,De Ruddere and Craig, 2016). The majority of the included participants were currently on sick-leave or early retirement due their CMP. The current results may therefore not be generalizable to people with CMP, who are trying to maintain their work status. The Danish healthcare system provides the possibility to receive economic compensation for one’s salary when on sick leave, and thus none of the participants mentioned economy as being a challenge.

### 4.6 Future research

The findings of the current study should inform the development and evaluation of future self-management interventions, in which it should be examined how the identified barriers and facilitators best possibly can be addressed to accommodate the daily challenges experienced by people living with CMP. This can potentially be done by developing theory-driven interventions, where the barriers and facilitators are implemented as specific treatment targets, and post-intervention analysis conducted to examine potential changes in these factors. Further, in our analysis many of the participants described self-management, acceptance and managing their daily resources as being a complex, dynamic and iterative learning process. Due to the current study only including interviews at a single timepoint, an in-depth understanding of self-management as a temporal learning process, with potential different needs for self-management support throughout, are yet to be uncovered. Therefore, researchers within the field of self-management and CMP should in future studies consider conducting prospective and longitudinal qualitative studies to unravel the learning process of self-management that people with CMP are going through, as well as the potential varying barriers for self-management, and needs for self-management support, throughout the different phases of this learning process.

## 5. Conclusion

This study identified several barriers and facilitators experienced by people with CMP during their everyday lives. These barriers and facilitators may inform clinicians currently providing self-management support as well as researchers trying to develop self-management interventions. Integrating the identified barriers and facilitators as specific treatment targets in the development of future self-management interventions might have the potential to improve the outcome of these interventions by taking a person-centered approach that addresses the specific biopsychosocial domains of importance to people living with CMP.

## Supporting information

Appendix files including 1- interview guide and 2- thematic map

## Data Availability

The data obtained during this study contains person sensitive information and will not be made publicly available to shield participants identities and ensure compliance with the EUGDPR

## Clinical protocols

No clinical protocols were uploaded to protocol repositories prior to the initiation of this study.

## Acknowledgements

The authors are grateful to the study participants, who agreed and were willing to spend their time and share their stories and experiences during the interviews. Furthermore, the authors would like to thank the employees from the rehabilitation centers in the Northern Denmark Region, who contributed throughout the recruitment process.

