## Appendix files including 1- interview guide and 2- thematic map for "Exploring Self-Management Barriers and Facilitators Experience by People with Chronic Musculoskeletal Pain - A Qualitative Study"

### Appendix 1 – Interview guide

| Research questions | Interview questions | Prompting questions (Optional) |
|---|---|---|
| <u>Theme 1: Course of illness</u><br>1) How has the participant course of illness been from onset until today? (Timeline exercise) | 1) Could you describe your history from the onset of your pain until today? | ● When did it start?<br>● Has there been periods during your history where it have been difficult to self-manage?<br>● Has there been periods during your history where it have been easier to self-manage?<br>● On a scale from 0 to 10 – How difficult was it to self-manage in that situation/period?<br>● On a scale from 0 to 10 – How easy was it to self-manage in that situation/period? |
| <u>Theme 2: Barriers for self-management</u><br>1) Which barriers does the participant experience, when trying to self-manage during everyday life? | 1) Could you describe some of the situations/periods where it was difficult for you to self-manage your condition?<br>2) What have you experienced as being the most difficult to self-manage? | ● Can you elaborate?<br>● How was that experience?<br>● Why was it difficult?<br>● What did it do to you?<br>● Have you experienced other situations/periods where it was difficult to self-manage? |

| | | |
|---|---|---|
| Theme 3: Facilitators for self-management<br>1) Which facilitators does the participant experience, when trying to self-manage during everyday life? | 1) What have helped you to self-manage the situations/periods, which were difficult for you to manage?<br>2) What have you experienced as being the greatest help to manage difficult situations/periods during you course of illness? | ● Can you elaborate?<br>● How was that experience?<br>● Why did it help you?<br>● Have you experienced other situations/periods, where you perceived something to be helpful for your ability to self-manage difficult situations/periods? |
| Theme 4: Completion<br>1) Does the participant experience other barriers or facilitators than those, we've currently addressed? | 1) Is there any other situations or periods during your course of illness, which have made it either more difficult or easier for you to self-manage you conditions? | ● Can you elaborate?<br>● What did you experience as being difficult?<br>● Why was that difficult for you?<br>● What did you experience as being helpful?<br>● Why was that helpful for you? |

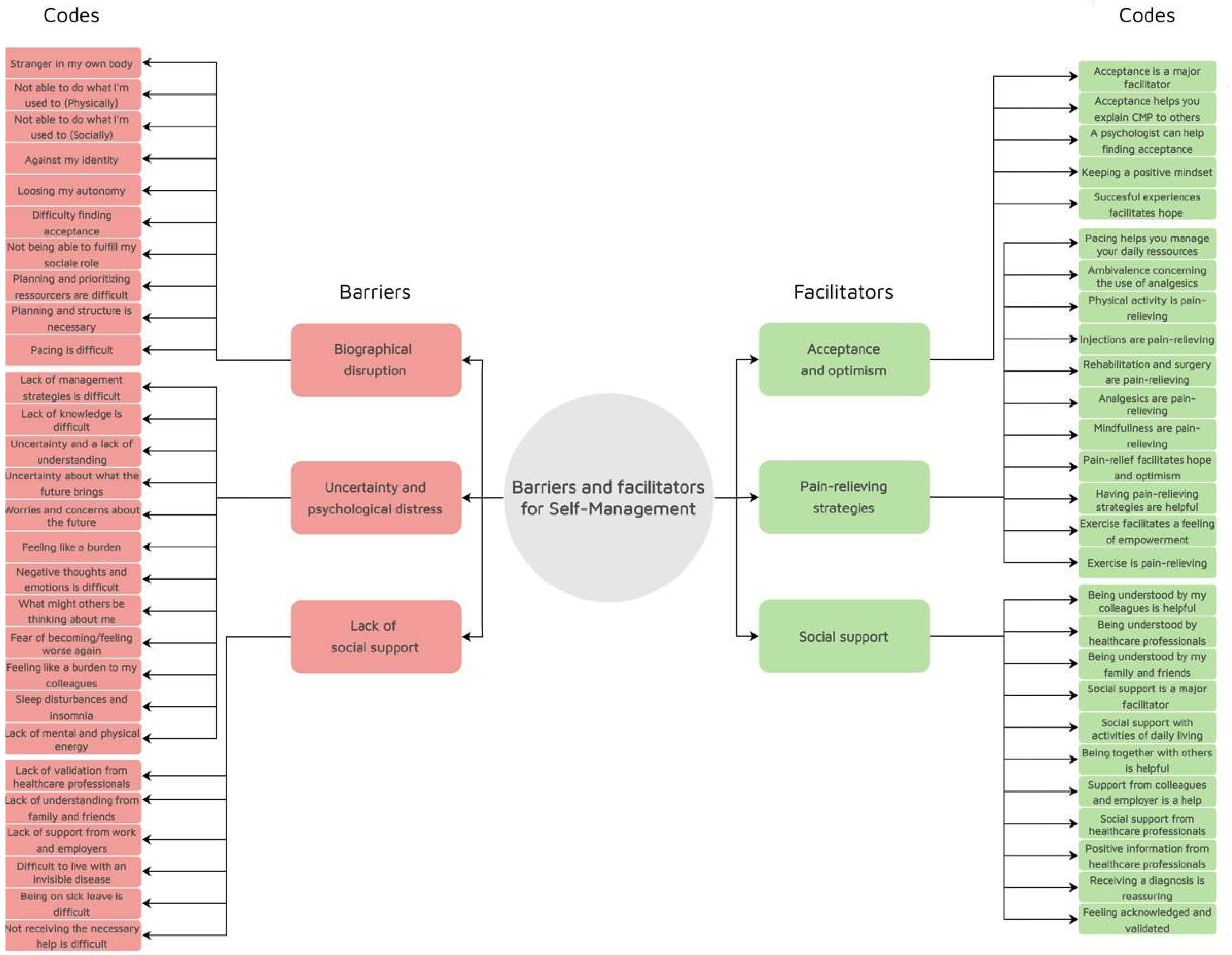

Codes
Codes
Stranger in my own body
Not able to do what I'm used to (Physically)
Not able to do what I'm used to (Socially)
Against my identity
Loosing my autonomy
Difficulty finding acceptance
Not being able to fulfill my sociale role
Planning and prioritizing ressourcers are difficult
Planning and structure is necessary
Pacing is difficult
Lack of management strategies is difficult
Lack of knowledge is difficult
Uncertainty and a lack of understanding
Uncertainty about what the future brings
Worries and concerns about the future
Feeling like a burden
Negative thoughts and emotions is difficult
What might others be thinking about me
Fear of becoming/feeling worse again
Feeling like a burden to my colleagues
Sleep disturbances and insomnia
Lack of mental and physical energy
Lack of validation from healthcare professionals
Lack of understanding from family and friends
Lack of support from work and employers
Difficult to live with an invisible disease
Being on sick leave is difficult
Not receiving the necessary help is difficult
Barriers
Biographical disruption
Uncertainty and psychological distress
Lack of social support
Barriers and facilitators for Self-Management
Facilitators
Acceptance and optimism
Pain-relieving strategies
Social support
Acceptance is a major facilitator
Acceptance helps you explain CMP to others
A psychologist can help finding acceptance
Keeping a positive mindset
Succesful experiences facilitates hope
Pacing helps you manage your daily ressources
Ambivalence concerning the use of analgesics
Physical activity is pain-relieving
Injections are pain-relieving
Rehabilitation and surgery are pain-relieving
Analgesics are pain-relieving
Mindfullness are pain-relieving
Pain-relief facilitates hope and optimism
Having pain-relieving strategies are helpful
Exercise facilitates a feeling of empowerment
Exercise is pain-relieving
Being understood by my colleagues is helpful
Being understood by healthcare professionals
Being understood by my family and friends
Social support is a major facilitator
Social support with activities of daily living
Being together with others is helpful
Support from colleagues and employer is a help
Social support from healthcare professionals
Positive information from healthcare professionals
Receiving a diagnosis is reassuring
Feeling acknowledged and validated
